# Science Map of Cochrane Systematic Reviews Receiving the Most Altmetric Attention: Network Visualization and Machine Learning Perspective

**DOI:** 10.1101/19006817

**Authors:** Jafar Kolahi, Saber Khazaei, Elham Bidram, Roya Kelishadi, Pedram Iranmanesh, Mohammad H. Nekoofar, Paul M. H. Dummer

## Abstract

We aimed to analyze and visualize the science map of Cochrane systematic reviews (CSR) with high Altmetric attention scores (AAS). On 10 May 2019, the Altmetric data of the CSR Database were obtained from the Altmetric database (Altmetric LLP, London, UK). Bibliometric data of the top 5% of CSR were extracted from the Web of Science. Keyword co-occurrence, co-authorship, and co-citation network analysis were then employed using VOSviewer software. A Random forest model was used to analyze the citation patterns. A total of 12016 CSR with AAS were found (Total mentions: 259968) with Twitter being the most popular Altmetric resource. Consequently, the top 5% (607 articles, mean AAS: 171.2, 95% confidence level (CL): 14.4, mean citations: 42.1, 95%CL: 1.3) with the highest AAS were included in the study. Keyword co-occurrence network analysis revealed female, adult, and child as the most popular keywords. Helen V. Worthington (University of Manchester, Manchester, UK), and the University of Oxford and UK had the greatest impact on the network at the author, organization and country levels respectively. The co-citation network analysis revealed that The Lancet and CSR database had the most influence on the network. However, AAS were not correlated with citations (r=0.15) although they were correlated with policy document mentions (r=0.61). The results of random forest model confirmed the importance of policy document mentions. Despite the popularity of CSR in the Twittersphere, disappointingly, they were rarely shared and discussed within the new academic tools that are emerging, such as F1000 prime, Publons, and PubPeer.

**Article Highlights:**

- The CSR database was most mentioned in Twitter.
- Twitter and News act as the greatest prominent issues regarding altmetric scores.

## Introduction

Cochrane is a British charity founded in 1993 by Iain Chalmers. The organization was created specifically to manage findings from medical research in order to facilitate evidence-based choices in health interventions faced by health professionals, patients, health policy makers, as well as those interested in health to make informed decisions about health promotion. Cochrane includes 53 review groups from 130 countries (Morley et al. 2016).

Alternative metrics, abbreviated to altmetrics, is an emerging academic tool that measures the online attention surrounding scientific research outputs (Konkiel 2017; Baheti and Bhargava 2017; Kolahi 2015). It complements, but does not replace, traditional citation-based metrics (Butler et al. 2017). Altmetric data resources include Twitter, Facebook (mentions on public pages only), Google+, Wikipedia, news stories, scientific blogs, policy documents, patents, post-publication peer reviews (Faculty of 1,000 Prime, PubPeer), Weibo, Reddit, Pinterest, YouTube, online reference managers (Mendeley and CiteULike), and sites running Stack Exchange (Q&A) (Patthi et al. 2017). In comparison with traditional citation-based metrics, altmetric data resources are updated rapidly. A recent bibliometric analysis revealed that only 50% of articles were cited in the first three years after publication (Wang 2013). In contrast, several altmetric data resources are updated in real-time (e.g. Twitter and Wikipedia) or on a daily-basis (e.g. Facebook).

Research funders and charities, such as the Wellcome Trust and John Templeton Foundation pay attention to altmetric analysis (Dinsmore et al. 2014). A study on the influence of the alcohol industry on alcohol policy would be a good example of their engagement (McCambridge et al. 2013). The study was supported by the Wellcome Trust, which invests approximately £600 million (US$ 936 million) a year in research, and alleged that several submissions to the Scottish government misrepresented research outputs so as to support policies preferred by the alcohol industry. Three months following this publication in PLOS medicine, it remained without citation. Yet, altmetrics allowed the Wellcome Trust to understand that this article had been tweeted by the key influencers, including members of the European Parliament, international nongovernmental organizations, and a sector manager for Health, Nutrition, and Population at the World Bank to reveal its global impact on the policy sphere (Dinsmore et al. 2014).

In the context of growing request among the research community to publicize research findings in cyberspace, new terms have appeared in the scientific literature, e.g. Twitter science stars (You 2014) and Kardashian index (a statistic measuring the over/under activity of scientists in the Twittersphere) (Hall 2014).

Altmetric research is a growing field in medical science (Azer and Azer 2019; Barbic et al. 2016; Kolahi and Khazaei 2016; Kolahi et al. 2017; Kolahi and Khazaei 2018; Kolahi et al. 2019b; Rosenkrantz et al. 2017; Wang et al. 2017). Here we aimed to analyze and visualize the knowledge structure of Cochrane systematic reviews with high altmetric attention scores to discover hot topics and influential researchers and institutions.

## Methods

On 10 May 2019, the Altmetric data associated with the Cochrane Database of Systematic Reviews were obtained from the Altmetric database (Altmetric LLP, London, UK). The bibliometric data of the top 5% Cochrane systematic reviews with the highest altmetric score were extracted from the Web of Science. Keyword co-occurrence, co-authorship, and co-citation network analysis were then employed using VOSviewer software (http://www.vosviewer.com/, Centre for Science and Technology Studies, Leiden University, Leiden, The Netherlands) (Van Eck and Waltman 2010).

The Pearson correlation coefficient was also used for the correlation analysis. In this step, citation counts (according to the Dimensions database (Van Noorden 2018)) and number of Mendeley readers were included along with altmetric data. A random forest model (a machine learning algorithm) was used to rank influential factors impacting on altmetric scores and citations. Random forest is a method of regression that creates a set of decision trees that consists of a great number of separate trees which operate as a group as like as a forest. Rattle 5.2.0 (https://rattle.togaware.com/, graphical user interface for data science in R) was used for data analysis (Williams 2011).

## Results

A total of 12016 Cochrane systematic reviews with altmetric attention were found (total mentions: 259,968). Twitter was the most popular altmetric resource among these articles (Figure 1) with Tweets originating mainly from the UK (18.8%), Spain (9.8%), and the US (7.8%).

**Figure 1.**
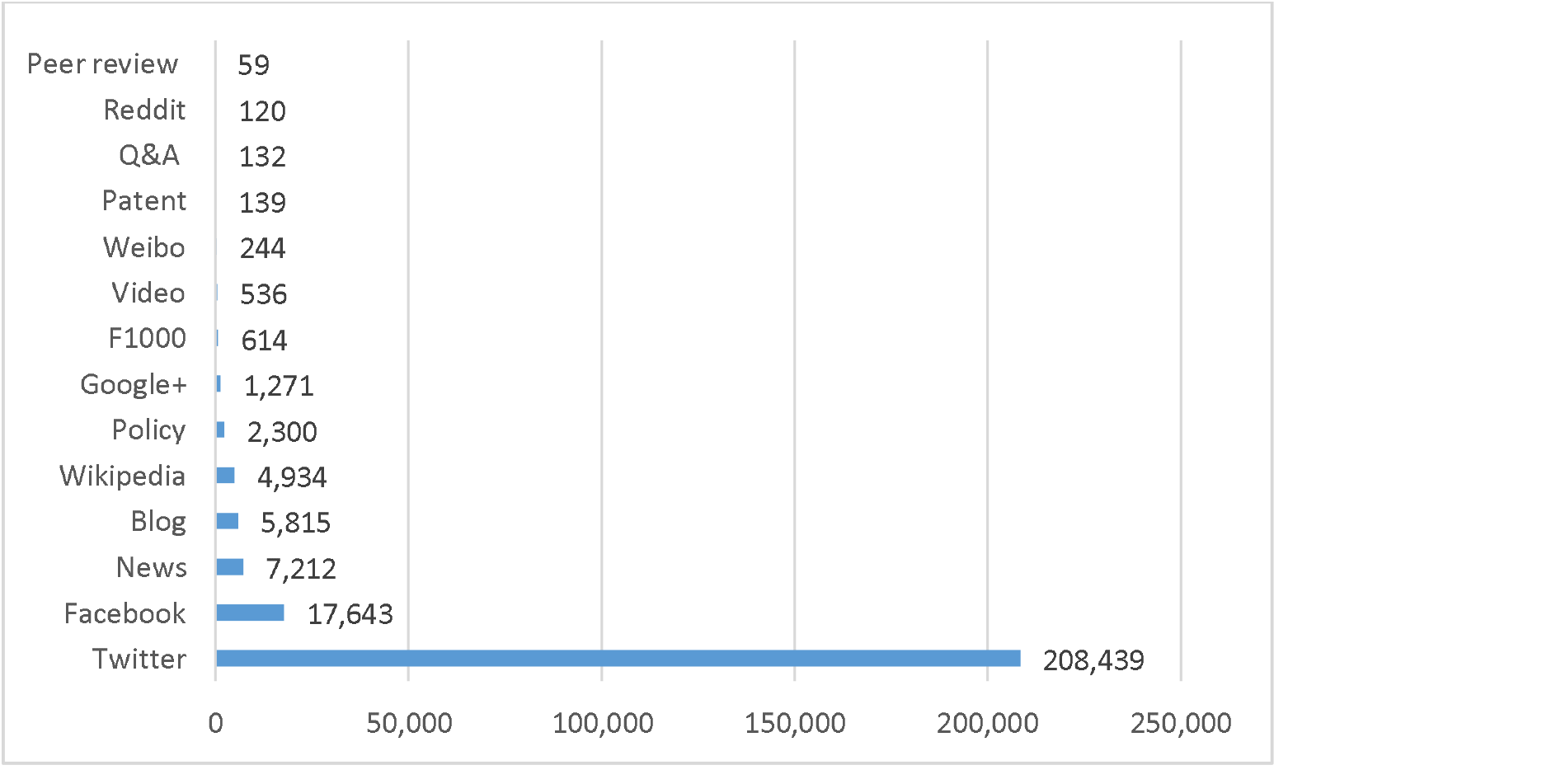
Sum of scores of various Altmetric data resources among all Cochrane systematic reviews.

Consequently, the top 5% (607 articles, mean altmetric score: 171.2, 95%CL: 14.4, mean citations: 42.1, 95%CL: 1.3) with the highest altmetric scores were included in the study. The bibliometric data of 552 articles found in the Web of Science were further analyzed. “Vitamin C for preventing and treating the common cold”, for instance, was a Cochrane systematic review with the highest altmetric score. This article was among the top 5% of all research outputs scored by altmetrics (Altmetric score: 587). It was discussed in 54 mainstream news outlets, 3 scientific blogs, 174 tweets (with an upper bound of 5,126,827 followers, in which 89% were made by members of the public), 91 Facebook pages, 4 Google+ pages, 1 Wikipedia article, 1 research highlight platform, 1 video up-loader, used by 754 Mendeley readers, and 2 CiteULike users (www.altmetric.com/details/1229303)(Table 1).

**Table 1.**
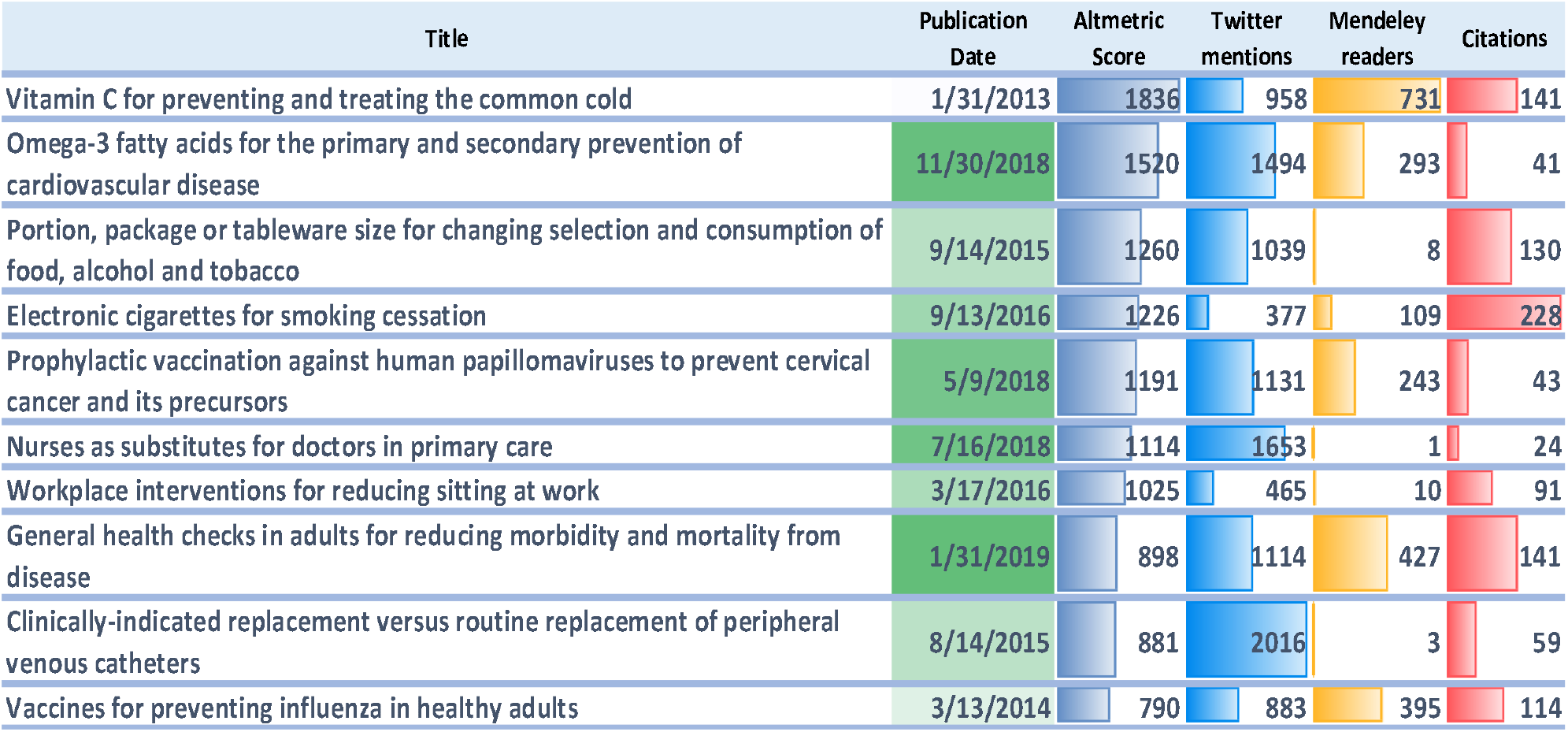
Data-bar visualization of the top ten Cochrane systematic reviews receiving the most altmetric attention

@CochraneUK (4,087 total mentions from this Twitter user) was the most active altmetric resource, followed by Cochrane zdravlje (1,702 total mentions from this Facebook wall) and @silvervalleydoc (1,662 total mentions from this Twitter user).

Keyword co-occurrence network analysis revealed that female, adult, and child were the most popular keywords (Figure 2). At author level, Helen V Worthington (Co-Coordinating Editor of the International Cochrane Oral Health Group) had the greatest impact on the network and Lee Hooper (Research Synthesis, Nutrition & Hydration at Norwich Medical School) had a central connection role in the network (Figure 3). At organization and country levels, University of Oxford and UK had the greatest impact on the network (Figure 4 and 5). Co-citation network analysis revealed that the Lancet and Cochrane Database of Systematic Reviews had the most influence on the network (Figure 6).

**Figure 2.**
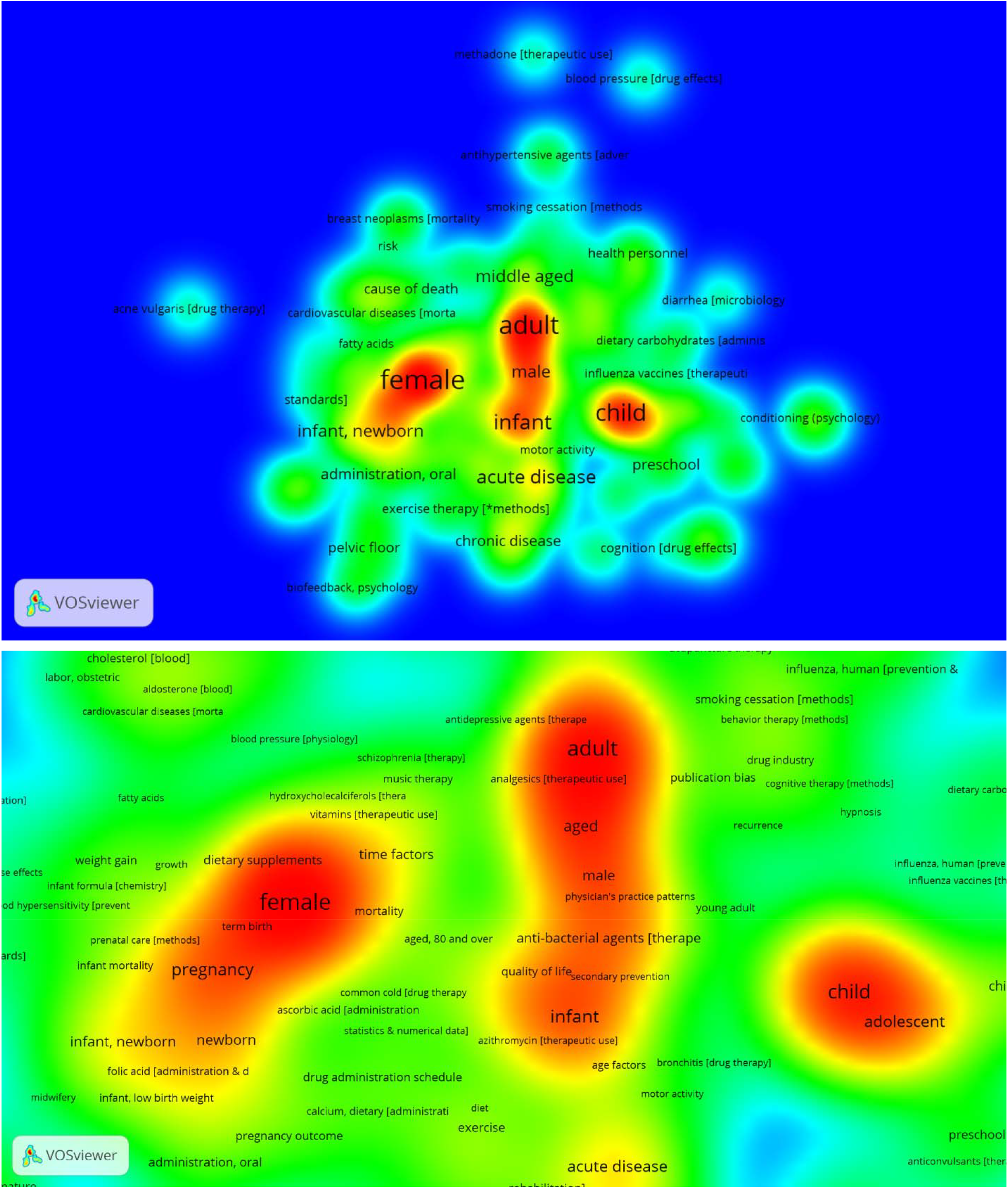
Hot topics among author keywords of the top 5% Cochrane systematic reviews receiving the most altmetric attention. The lower part zoomed on central hot zones. The distance-based approach was used to create this map, which means the smaller distance between two terms, the higher their relatedness.

**Figure 3.**
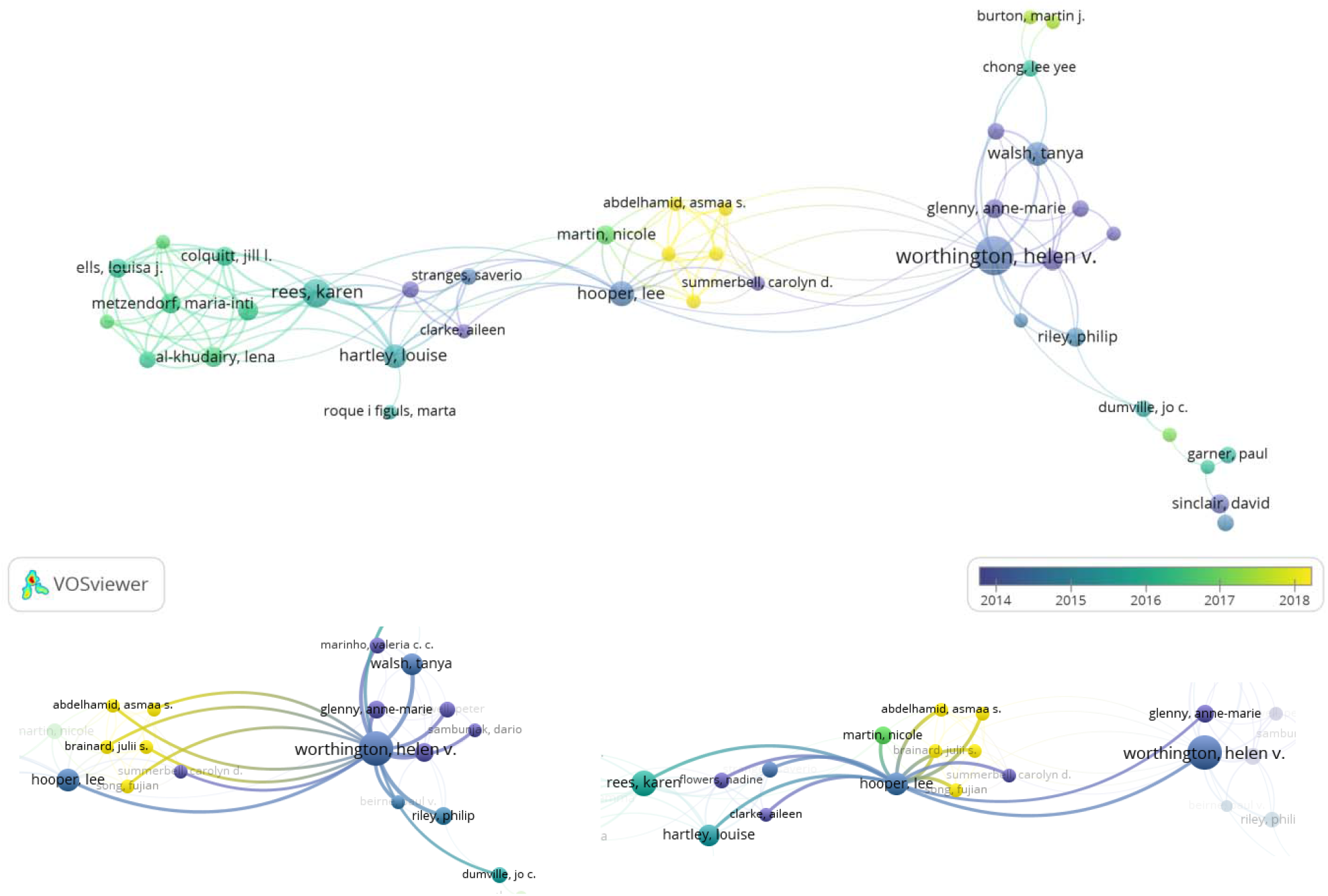
Co-authorship network visualization of top 5% Cochrane systematic reviews receiving the most altmetric attention. Lower part showed personal networks of Helen V Worthington and Lee Hooper. These two people had deep connections with contemporary growing influential authors (yellow nodes).

**Figure 4.**
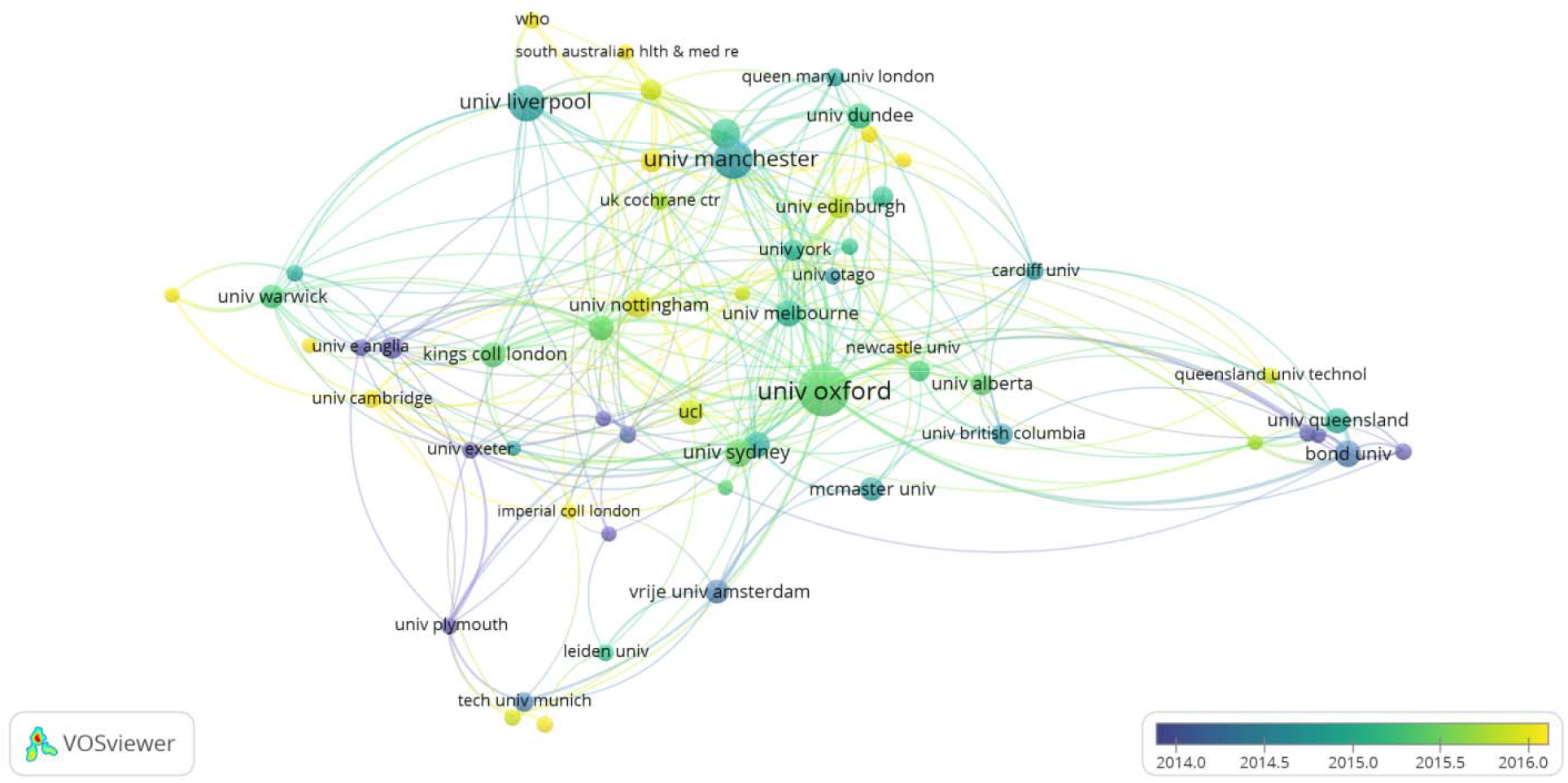
Organization level co-authorship network visualization of top 5% Cochrane systematic reviews receiving the most altmetric attention.

**Figure 5.**
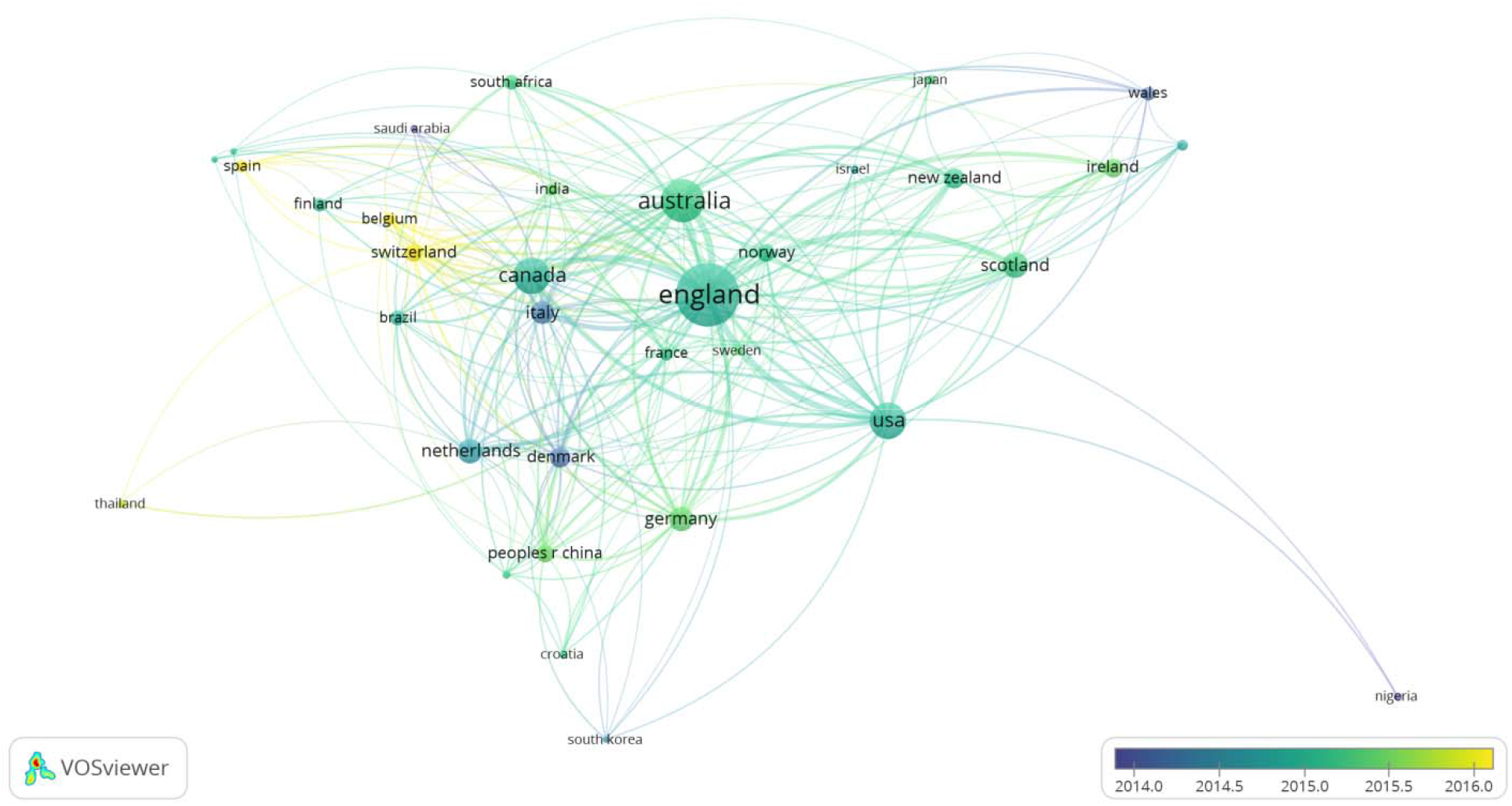
Country level co-authorship network visualization of top 5% Cochrane systematic reviews receiving the most altmetric attention.

**Figure 6.**
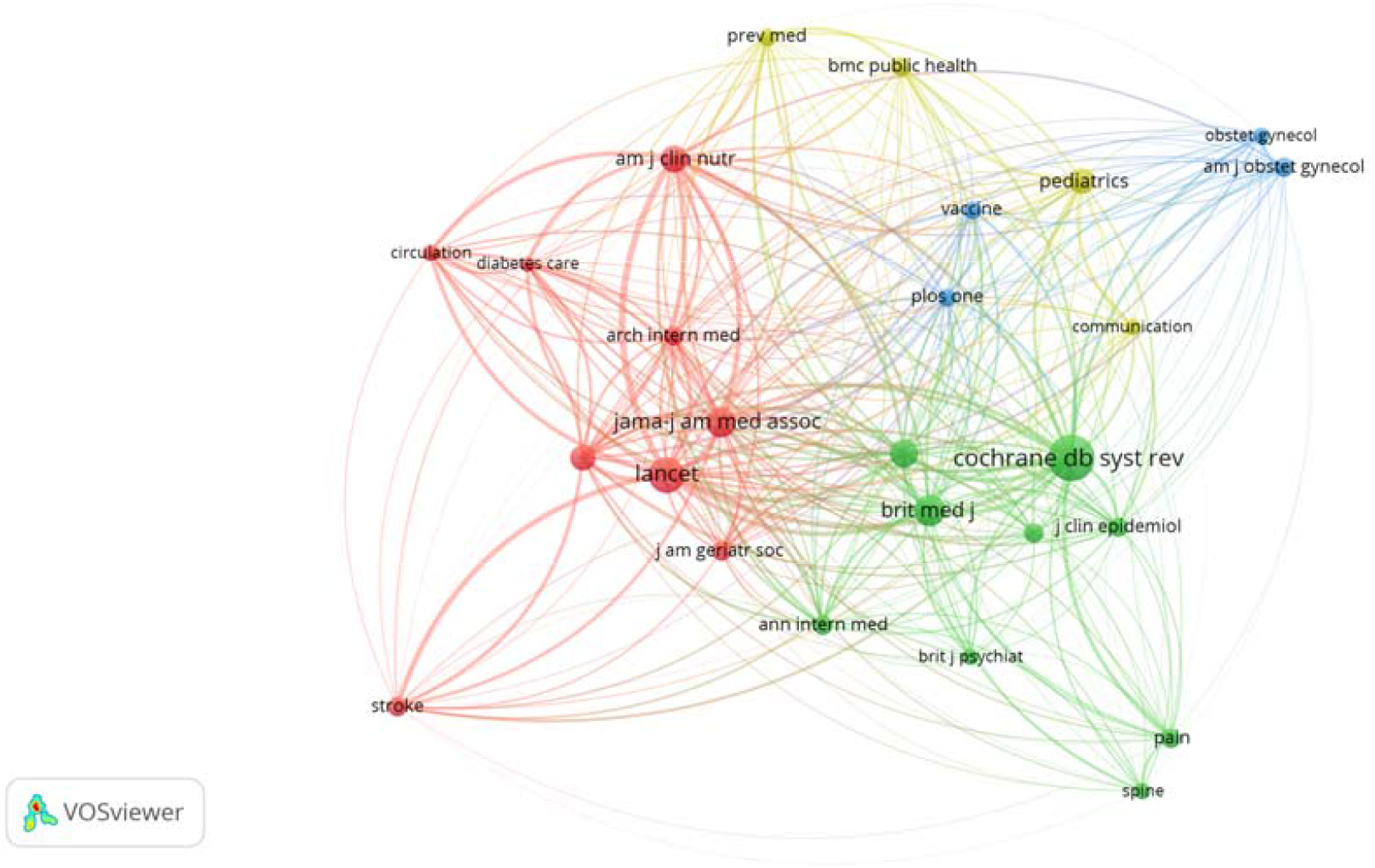
Co-citation network visualization among resources of top 5% Cochrane systematic reviews receiving the most altmetric attention.

Altmetric scores were not correlated with citations (r=0.15) (Figure 7 and 8) but were correlated with policy document mentions (r=0.61) (Figure 7). Results from the random forest model confirmed the importance of policy document mentions regarding citations. Twitter and News were the most influential factors regarding altmetric scores (Figure 9).

**Figure 7.**
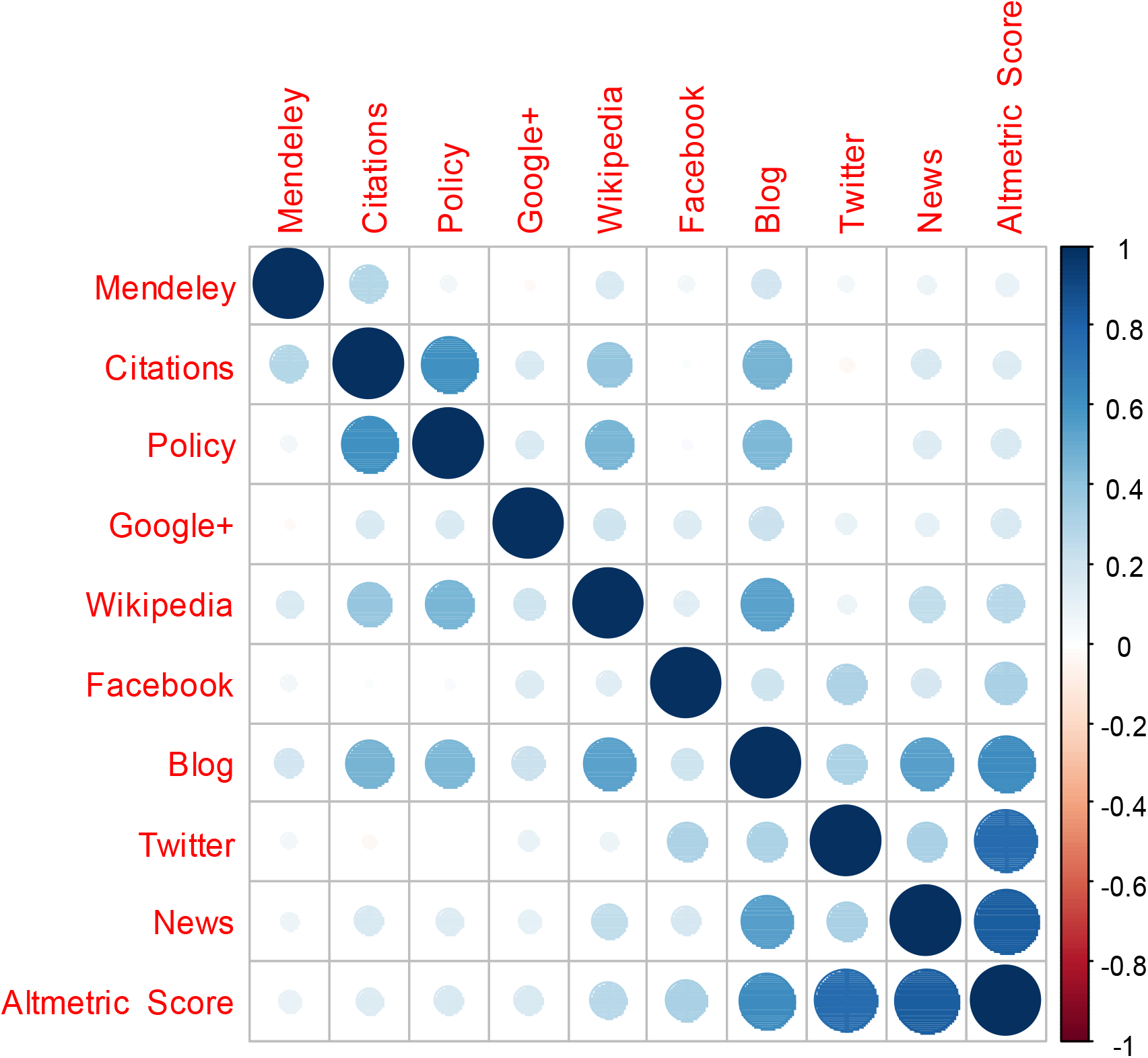
Correlation matrix visualization between Altmetric score, citations, Mendeley readers and other important altmetric resources.

**Figure 8.**
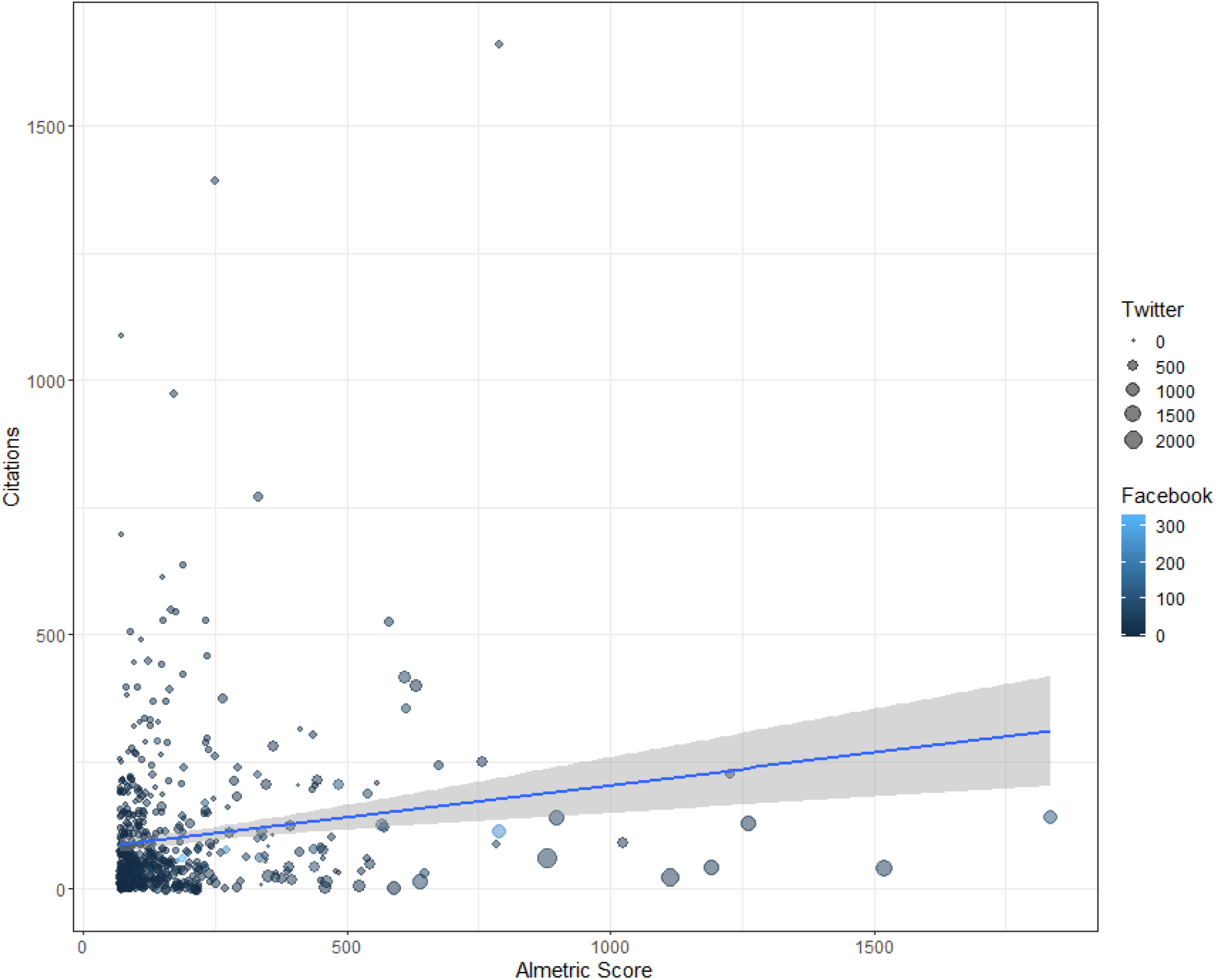
Scatter plot examining the relationship between Altmetric score and citations. Fitted line represents the linear regression model with 95% confidence interval.

**Figure 9.**
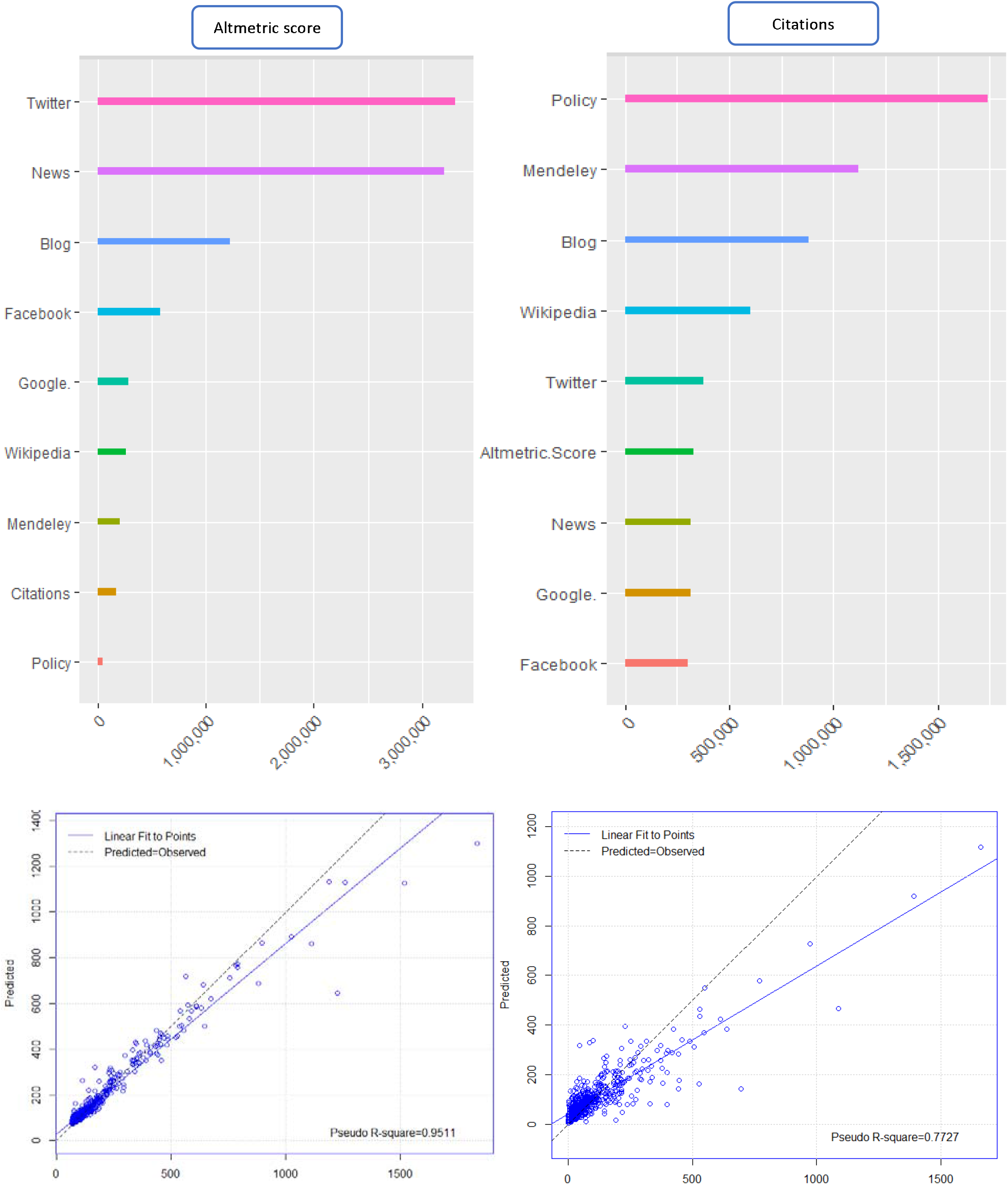
Output of random forest model showing relative importance of different altmetric resources considering Altmetric score (left side) and citations (right side) as targets (Number of observations used to build the model: 424, Number of trees: 500). Accuracy of the model showed in the lower part via predicted versus observed plot.

## Discussion

It has been estimated that the number of social media users will increase to 3.09 billion by 2021(Clement 2019). A large-scale survey revealed that Twitter plays a key role in the discovery of academic information (Mohammadi et al. 2018) and that well-known academic healthcare providers use social media to communicate with patients and disseminate trusted medical information (Pershad et al. 2018). For example, @MayoClinic with 1.92 million followers and 47600 tweets (Widmer et al. 2016). It is widely believed that Cochrane systematic reviews are one of the most important resources in evidence-based clinical decision making. In the present study, we analyzed the social impact of this reliable medical evidence using altmetrics (Dinsmore et al. 2014).

Overall, analysis of the top 5% of Cochrane systematic reviews had high altmetric scores (mean =171.2). Twitter was the most popular resource with tweets generally originating from the UK. Popular Cochrane systematic reviews received a high citation rate (mean: 42.1), however, there was no significant correlation between citations and altmetric score. Likewise, no significant correlation was reported between citations and altmetric score for original research articles published in high-impact general medicine journals (Barakat et al. 2019), the cardiovascular field (Barakat et al. 2018), dentistry (Delli et al. 2017), radiology (Rosenkrantz et al. 2017), and Iranian medical journals (Kolahi et al. 2019a). In contrast, a large-scale survey indicated significant correlation between six altmetric resources (tweets, Facebook wall posts, research highlights, blog mentions, mainstream media mentions, and forum posts) and citation counts (Thelwall et al. 2013). Among six PloS specialized journals, a significantly positive correlation was found between the normalized altmetric scores and normalized citations (Huang et al. 2018). In the field of general and internal medicine, the number of Twitter followers was significantly associated with citations and impact factor (Cosco 2015). However, correlation between altmetric score and citations is a conraversal issue and more studies needed to solve this contaversy.

Despite the popularity of Cochrane systematic reviews in Twittersphere, they were rarely shared and discussed within the emerging academic tools, such as F1000 prime, Publons and PubPeer. Although English medical Wikipedia articles received more than 2.4 billion official visits in 2017 (Murray 2019) the overall Wikipedia mentions among Cochrane systematic reviews was low considering the established partnership between Wikipedia and Cochrane (Dawson 2020).

The keyword co-occurrence network analysis revealed that newly emerging and ground-breaking concepts, e.g. genomic medicine, nanotechnology, artificial intelligence were not included among the hot topics. This finding may be explained by retrospective nature of Cochrane systematic reviews.

Helen V Worthington (Coordinating Editor of the International Cochrane Oral Health Group) had the greatest impact on the network based on co-authorship network analysis. The results of PubMed query *Worthington, Hv [Full Author Name] OR Worthington HV [Author]* revealed142 Cochrane systematic reviews, from which nine of these articles were withdrawn. Another influential author, Lee Hooper (Research Synthesis, Nutrition & Hydration at Norwich Medical School), had three withdrawn articles among 26 Cochrane systematic reviews. Further examination with the PubMed query *“The Cochrane database of systematic reviews”[Journal] AND WITHDRAWN[Title]* revealed 467 withdrawn Cochrane systematic reviews. Article withdrawal is defined as taking back an article from a journal, either by author or by editor just before final publishing or article in press status. According to a Cochrane editorial and publishing policy resource, Cochrane Reviews should only be withdrawn under exceptional circumstances, i.e. serious error, serious breach of Cochrane’s conflict of interest policy and scientific misconduct in the Cochrane Review (MacLehose 2019).

It has been reported that journals with their own Twitter account get 34 percent more citations and 46 percent more tweets than journals without a Twitter account (Ortega 2017). Of more interest, the Cochrane organization and the related groups such as @cochranecollab (83.9K followers and 11.9K Tweets), @CochraneUK (46.6K followers and 31.1K Tweets), @CochraneLibrary (58.8K followers and 5.9K Tweets), @CochraneCanada (5K followers and 5.2K Tweets) were quite active in the Twittersphere. Among the 12016 Cochrane systematic reviews included in this study, the total number of citations was 506,100. With respect to the ratio of number of tweets (≈54100) to the number of citations (506,100), it was estimated that Cochrane was not a Kardashian organization (overactive in Twittersphere) (Hall 2014).

## Data Availability

Data is available. Please contact with

## Acknowledgment

We would like to thank Altmetric LLP (London, U.K), particularly Mrs. Stacy Konkiel for her valued support and permitting us complete access to Altmetric data.

